# Drug classes affecting intracranial aneurysm risk: genetic correlation and Mendelian randomization

**DOI:** 10.1101/2023.10.23.23297443

**Authors:** Ynte M. Ruigrok, Jan H. Veldink, Mark K. Bakker

**Author notes:** **Correspondence** Mark K. Bakker, Department of Neurology, University Medical Center Utrecht Brain Center, University Medical Center Utrecht, The Netherlands. Heidelberglaan 100, 3584 CX Utrecht, The Netherlands, Twitter: @bakker_mk.

## Abstract

**Background:** There is no non-invasive treatment option to prevent aneurysmal subarachnoid hemorrhage (ASAH) caused by intracranial aneurysm (IA) rupture. We aimed to identify drug classes that may affect liability to IA using a genetic approach.

**Methods:** We obtained genome-wide association summary statistics for unruptured IA (N=2,140 cases), ASAH (N=5,140 cases), and the combined group (N=7,495 cases; N=71,934 controls in all groups), and drug usage from 23 drug classes (N up to 320,000) based on European-ancestry cohorts. We calculated genetic correlation between IA and ASAH, and liability to drug usage independent of the risk factor high blood pressure. Next, we evaluated the causality and therapeutic potential of independent correlated drug classes using three different Mendelian randomization frameworks.

**Results:** Blood pressure-independent correlations with IA were found for antidepressants, paracetamol, acetylsalicylic acid, opioids, beta-blockers, and peptic ulcer and gastro-oesophageal reflux disease drugs. MR showed that the genetically predicted usage of none of these drug classes were causally related to IA. Genetically predicted high responders to antidepressant drugs were at higher risk of IA (odds ratio [OR]=1.61, 95% confidence interval (CI)=1.09-2.39, P=0.018) and ASAH (OR=1.68, 95%CI=1.07-2.65, P=0.024) if they used antidepressant drugs. This effect was absent in non-users. For beta-blockers, additional analyses showed that this effect was not independent of blood pressure after all. Genetic liability to chronic multisite pain, an indication for pain medication (paracetamol, acetylsalicylic acid, and opioids), was associated with increased IA risk (OR=1.63, 95%CI=1.24-2.14), but not consistently across sensitivity analyses.

**Conclusions:** We did not find drugs decreasing liability to IA and ASAH but did find that antidepressant drugs may increase liability. In addition, we observed pleiotropy between IA, chronic pain, and pain medication usage, but the driving factor for this pleiotropy remains to be determined. Our results help to better understanding pathogenic mechanisms underlying IA.

## Introduction

Rupture of an intracranial aneurysm (IA) causes aneurysmal subarachnoid hemorrhage (ASAH), a severe stroke occurring typically at a relatively young age. ^1^ ASAH has devastating consequences and is responsible for a substantial economic burden. ^2^ ASAH can be prevented by treatment of an unruptured IA, but risk of procedural complications often outweighs the potential benefit of preventive treatment. ^3^ Therefore, safer options are needed to prevent ASAH.

There is currently no drug available that can prevent growth or rupture of an IA. Genetically informed drug targets are more likely to lead to approved drugs, ^4^ and usage of drugs within several drug classes was shown to be in part heritable. ^5^ Important risk factors for IA and ASAH are smoking and hypertension, ^6,7^ but genetic risk also plays a key role in IA development and in ASAH. ^8,9^ Therefore, IA may be a suitable disease to investigate drug targets for using a genetic approach. Previously we found overlap between IA and targets within the anti-epileptic drug class, ^10^ and later highlighted *CNNM2* as the main driver of that overlap using Mendelian randomization (MR). ^11^ We hypothesize that additional drug classes are present that can help to direct drug discovery. MR is an approach to assess causality of an exposure (in this study, drug usage) on an outcome (ruptured IA, unruptured IA, and the combined group) if specific assumptions are met. ^12^

We aimed to identify drug classes affecting the liability to IA to gain insight in the development and/or rupture of IA and potentially identify novel therapeutic mechanisms. For this we used a two-step genetic approach. First, we estimated the extend of genetic overlap between the liability to the usage of drugs within a drug class on the one hand, and IA (unruptured IA and ASAH as separate groups and combined) on the other hand. To identify novel mechanisms, we performed these analyses independent of blood pressure (BP). Next, for drug class correlated with IA independent of BP we leveraged genetic information to identify potential causal effects of usage of drug classes on IA liability, anticipate the effect of the use drugs within a class to treat IA, and assess the causality of the drug indication on IA liability, all using MR.

## Methods

In this study a combination of genetic techniques was used with an emphasis on MR. Therefore, we followed the STROBE-MR guidelines. ^13^ Only publicly available aggregated data was used with informed consent in place in the original studies. Data generated in this study is available in the Supplement. An overview of the methods pipeline is shown in Figure 1 and the outlines of these methods are described below. More details for all methods can be found in the supplementary Data.

**Figure 1.**
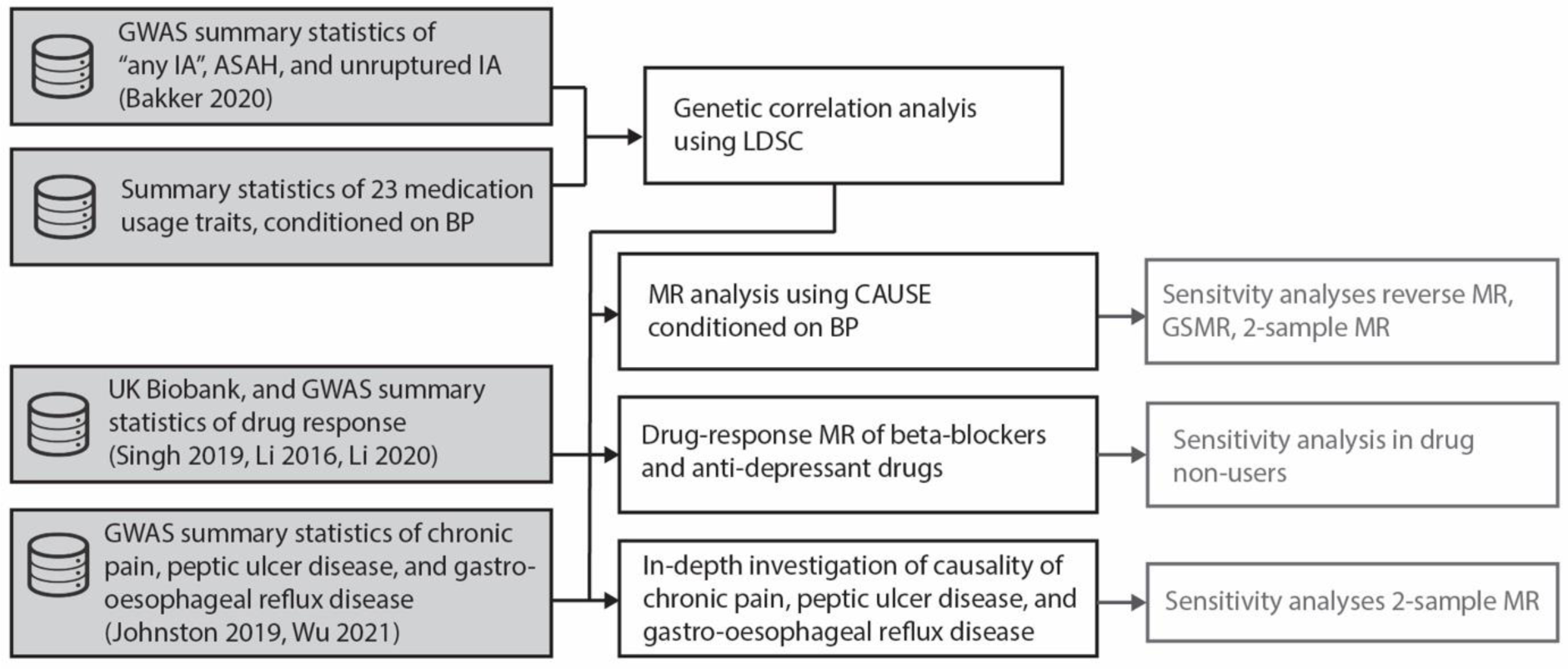
Overview of the methods. Analyses indicated in black boxes are part of the main analysis pipeline, while those in greyed-out boxes are sensitivity analyses. Input datasets are indicated in shaded boxes. IA: intracranial aneurysm. ASAH: aneurysmal subarachnoid hemorrhage. BP: blood pressure. GWAS: genome-wide association study. AnyIS: any ischemic stroke. LAS: large artery stroke. CES: cardioembolic stroke. SVS: small vessel stroke. ICH: intracerebral hemorrhage. LDSC: linkage disequilibrium score regression. CAUSE: Causal Analysis Using Summary Effect Estimates.

### Datasets

We obtained summary statistics of genome-wide association studies (GWAS) of 23 drug usage traits measured in the UK Biobank, ^5^ ASAH, unruptured IA, and IA (the combined group of ASAH and unruptured IA), ^10^ and BP (http://www.nealelab.is/uk-biobank). See Supplementary Table 1 for study details.

To exclude the role of BP, with increased BP being a main risk factor for IA, ^6,7^ we conditioned the summary statistics for the 23 drug usage traits on systolic and diastolic BP, using mtCOJO. This approach mimics the use of BP as a covariate in the source GWAS and allows the identification of IA-associated drug classes that present novel therapeutic mechanisms. ^14^

For drug response analyses we used the UK Biobank dataset. UK Biobank data are available to bona fide researchers on application at http://www.ukbiobank.ac.uk/using-the-resource/. When analyzing drug usage summary statistics in relation to summary statistics for ASAH, unruptured IA, and IA, we excluded UK Biobank samples in the IA analysis to avoid bias due to shared samples.

### Genetic correlation between drug usage and intracranial aneurysms

Genetic correlation was calculated between IA (unruptured IA, ASAH, and the combined group) and drug class usage (conditioned on BP) with LDSC. ^15^ Genetic correlation indicates the degree of shared heritability, having an estimate of ρ_g_=1 (or 100%) if traits are fully correlated, ρ_g_=0 if traits do not share heritability, and ρ_g_=−1 (or −100%) if traits are inversely correlated. ^16^ Trait pairs with a statistically significant genetic correlation (see Statistics paragraph in the Methods) were selected for subsequent MR analyses.

### Mendelian randomization analysis of drug usage on intracranial aneurysms

MR can infer causality of an exposure on an outcome if three assumptions are met: 1. the genetic variants used are associated with the exposure, 2. there are no unmeasured confounders between the exposure and the outcome, and 3. the genetic variants only affect the outcome through the exposure. ^12^ We defined usage of drug classes as exposure and IA (unruptured IA, ASAH, and the combined set) as outcome in our main analysis.

We selected Causal Analysis Using Summary Effect Estimates (CAUSE) as MR method since it models correlated and uncorrelated horizontal pleiotropy, thereby being more robust to reverse causality than other MR methods, and models the presence of unmeasured confounding factors. ^17^ We applied CAUSE to assess the effect of genetic liability for drug usage on the susceptibility to IA. In brief, CAUSE tests if a model that includes unmeasured confounders and a causal effect performs better than a model including only unmeasured confounders. In theory, since CAUSE can account for unmeasured confounders, conditioning drug usage summary statistics on BP therefore should not be necessary. However, as additional check we also performed the analysis using the summary statistics for drug usage conditioned on BP.

We performed the following sensitivity analyses for statistically significant MR effects identified with CAUSE: 1. reverse MR with IA as exposure and drug usage an outcome, and 2. other MR methods being generalized summary statistics-based MR (GSMR), ^14^ inverse variance weighted MR^18^, weighted mode MR, ^19^ and MR-Egger. ^20^

### Drug response Mendelian randomization

MR of drug usage risk on IA liability does not differentiate between causality due to the drug indication, drug usage, or other (pleiotropic) pathways. If a GWAS on the responsiveness to a drug is available, drug users can be divided based on their genetically predicted response. ^21^ Low responders closely resemble a placebo group, where the drug has no or lower effect, while participants are being aware of their responsiveness. We applied this framework to response GWASs that were available for drug classes that correlated with IA.

Using data from the UK Biobank, we extracted users of a drug class of interest (included drugs in Supplementary Table 2). We calculated a polygenetic score (PGS) for response to the drug class in those individuals. We then tested if having a top versus bottom tertile of PGS affected the risk of IA and ASAH, or ASAH hazard using logistic regressions and a Cox regression, respectively.

To confirm that the observed effect was due to usage of the drug class, we repeated the analysis in non-users of the drug to confirm absence of the PGS effect in non-users.

### Mendelian randomization analysis of indications for drug classes correlated with intracranial aneurysms

For some drug classes of which usage was correlated with IA, no drug response GWAS was available. To gain a better understanding of the observed genetic overlap of these drug classes with IA, we performed additional MR analyses using GWAS data of the indications of those drugs. We used the inverse variance weighted MR method implemented in R package TwoSampleMR to assess causality of drug indications on IA and its subtypes. We performed sensitivity analyses according to an existing framework. ^22^ We further tested if these diseases indeed were causal for the use of the respective drugs using the same MR approach.

### Statistics

Missing SNPs in either the exposure or outcome GWAS were excluded. We set the multiple testing threshold for genetic correlation and main MR analysis at 0.05/20=2.5×10^-3^, where 20 was the number of independent tests obtained according to the method described in the Supplementary Data (Supplementary Figures 1-4). All p-values were based on two-sided tests, except for the comparison of the sharing and causal models with CAUSE, which is a one-sided test.

## Results

### Genetic correlation between drug usage and intracranial aneurysms

We observed genetic correlations independent of BP between IA and usage of antidepressant drugs, anilides, salicylic acid and derivatives, opioid drugs, beta-blockers, and drugs for peptic ulcer and gastro-oesophageal reflux disease (Figure 2, Supplementary Table 3). The anilides drug class was completely comprised of paracetamol, while the salicyclic acid and derivatives class was completely comprised of acetylsalicylic acid (Supplementary Table 4). Therefore, from here on we refer to paracetamol and acetylsalicylic acid instead of the drug class names. Other highly represented drugs were omeprazole (approximately 50% of the peptic ulcer and gastro-oesophageal reflux disease drugs), and paracetamol and codeine in the opioid drugs class (66% and 54%, respectively). The antidepressant drug class was comprised of mostly selective serotonin reuptake inhibitors (approximately 50%) and tricyclic antidepressants (approximately 30%).

**Figure 2.**
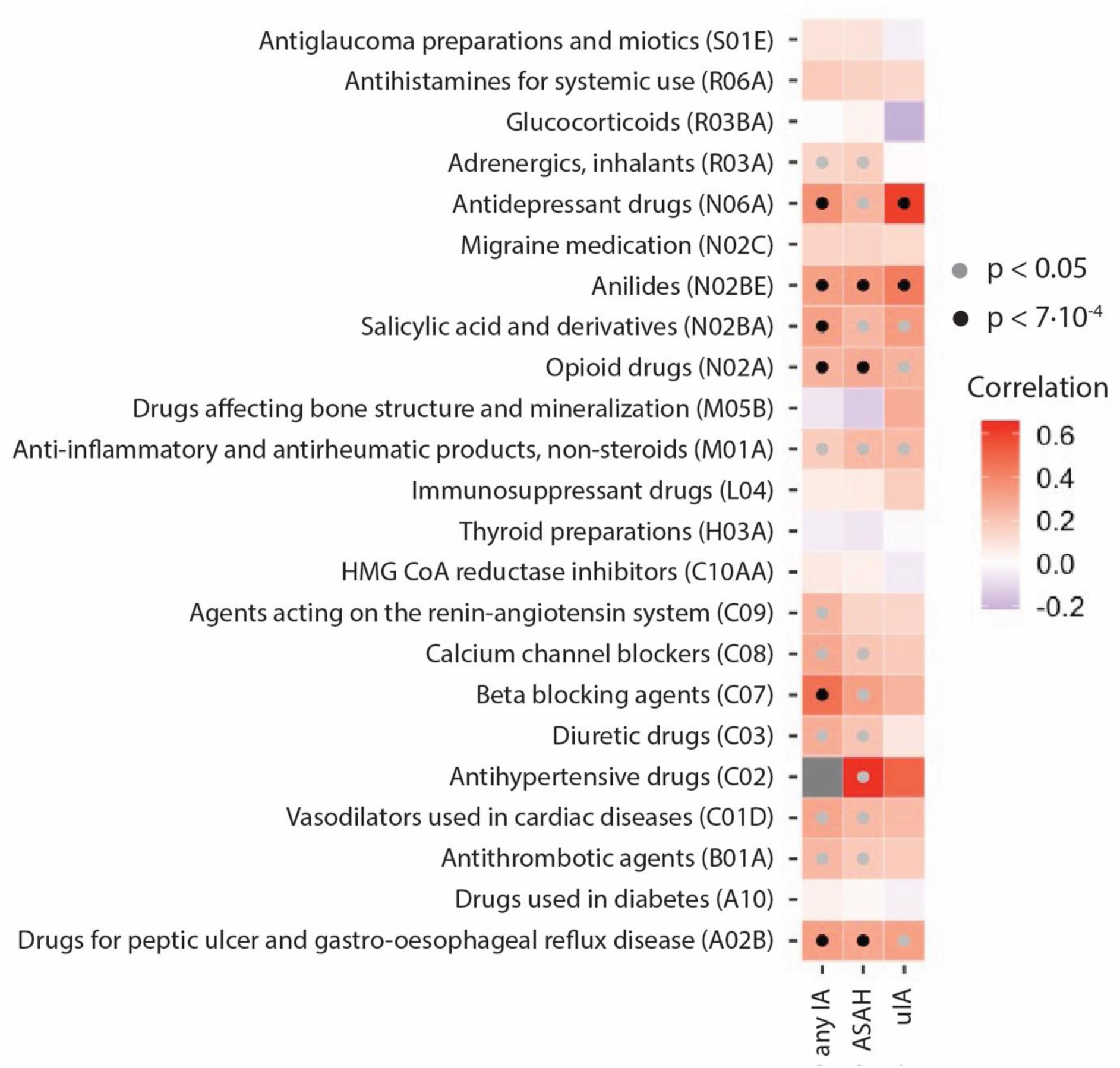
Genetic correlation analysis between intracranial aneurysms (IA), and drug usage conditioned on BP. Black dots indicate statistical significance of the genetic correlations (grey: nominally statistically significant, black: statistically significant after correction for multiple testing). *ASAH: aneurysmal subarachnoid hemorrhage. uIA: unruptured IA.

### Mendelian randomization analysis of drug usage on intracranial aneurysms

Of the drug classes that correlated with IA, only beta-blockers appeared to have a statistically significant MR effect (Figure 3A, Supplementary Table 5). Although CAUSE can account for unmeasured confounders, we aimed to confirm the independence of BP by using drug usage summary statistics conditioned on BP. Here, we found no evidence for a causal effect of beta-blockers usage on IA (Figure 3B). This indicates that CAUSE did not fully account for confounding by BP, and the correlation between IA and beta-blocker usage is driven by BP.

**Figure 3.**
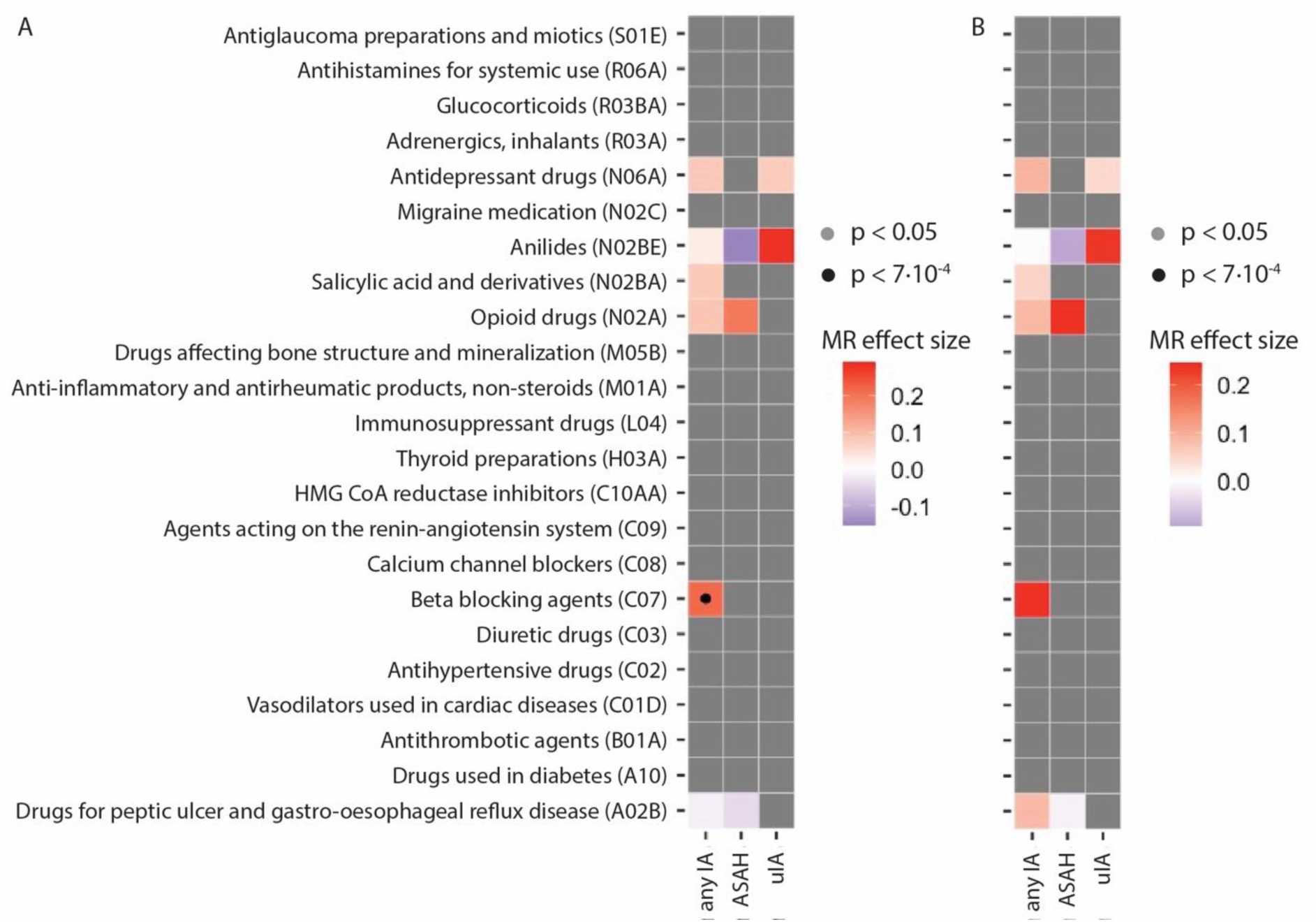
Mendelian randomization (MR) analysis of drug usage on liability to IA. A) Black dots indicate that a model that included a causal effect performs statistically significantly better than a model with only confounders at statistical significance after correcting for multiple testing. Colors indicate the MR effect estimate of the exposure (on the left) on the outcome (at the bottom). Grey boxes indicate exposure-outcome pairs that were not genetically correlated. B) MR estimates of an additional analysis with drug usage summary statistics conditioned on BP, showing that CAUSE did not fully account for BP as confounder when using beta-blockers as exposure.

Sensitivity analyses using additional MR algorithms showed the same directions of effect, and dependence on BP (Supplementary Table 6).

### Drug response Mendelian randomization

For the beta-blocker and antidepressant drug classes, drug response GWASs were available: blood pressure response to beta-blockers, ^23^ non-treatment-resistant depression (i.e., response to at least one anti-depressant drug), ^24^ response to selective serotonin reuptake inhibitors, ^24^ and response to antidepressant drugs citalopram and escitalopram. ^25^ Drug usage numbers are shown in Supplementary Table 4.

No difference in baseline characteristics were found between low and high predicted responders for each drug class (Supplementary Table 7). Of the four tested drug classes, genetically predicted higher response to antidepressants was associated with an increase in IA risk in antidepressant drug users (odds ratio [OR]=1.61, 95% CI=1.09-2.39, P=0.018, Figure 4, Supplementary Table 8). For response to SSRI’s and response to citalopram and escitalopram, large confidence intervals and no statistically significant effects were observed (SSRI: OR=1.19, 95% CI=0.66-2.12, P=0.56; citalopram/escitalopram: OR=0.98, 95% CI=0.47-2.07, P=0.97). The effect of genetically predicted antidepressant response on IA was absent in non-users of antidepressant drugs (OR=1.02, 95% CI=0.88-1.18, P=0.80).

**Figure 4.**
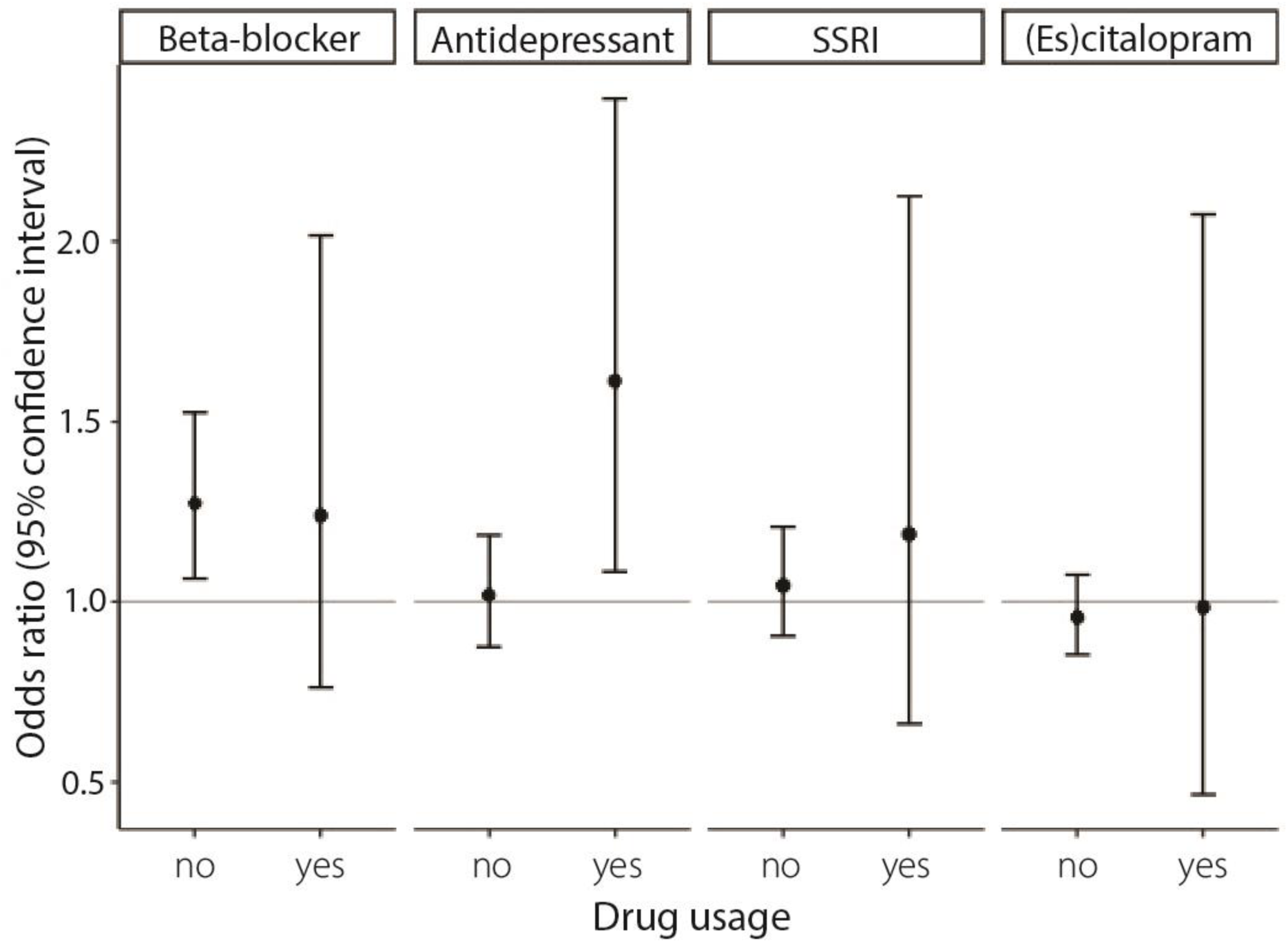
Drug response Mendelian randomization (MR) analysis. Genetically predicted response to beta-blockers, antidepressant drugs, selective serotonin reuptake inhibitor (SSRI) drugs, and citalopram/escitalopram were selected as exposures, and IA was selected as outcome. Per drug, the effect of high versus low predicted response was analyzed in drug users (the group of interest) and non-users (to rule out pleiotropy).

Beta-blocker response was associated with increased IA risk in non-users of beta-blockers (1.27, 95% CI=1.06-1.53, P=0.0085) and with similar effect size but large confidence intervals in beta-blocker users (1.24, 95% CI=0.76-2.03, P=0.39), indicating that genetically predicted beta-blocker response was associated with IA through another mechanism than through the effect of beta-blockers.

Highly similar effects throughout all drug classes were observed using ASAH as outcome, and in the Cox regressions for ASAH hazard (Supplementary Figures 5-6).

### Mendelian randomization analysis of indications for drug classes correlated with intracranial aneurysms

Since no drug response GWASs were available for peptic ulcer and gastro-oesophageal reflux disease drugs, opioid drugs, acetylsalicylic acid, and paracetamol, we performed MR with GWASs of indications for these drugs as exposure instead. Traits we included were peptic ulcer disease, ^26^ gastro-oesophageal reflux disease, ^26^ and chronic multisite pain (CMP). ^27^

CMP was consistent with a causal effect on IA using the inverse variance weighted approach (OR=1.63, 95% CI=1.24-2.14, P=4.7×10^-4^, Supplementary Figure 7, Supplementary Table 9). However, no statistically significant effects were found for most sensitivity analyses indicating a pleiotropic relationship between IA and CMP. We confirmed that genetic liability to CMP increased risk of paracetamol usage (Supplementary Table 9). We found no effect of genetic predisposition of peptic ulcer disease (OR=0.98, 95% CI=0.86-1.12, P=0.78) or gastro-oesophageal reflux disease (OR=1.13, 95% CI=0.96-1.32, P=0.13) on IA.

We summarized the observed relationship between IA, drug responses, and drug indications in Figure 5.

**Figure 5.**
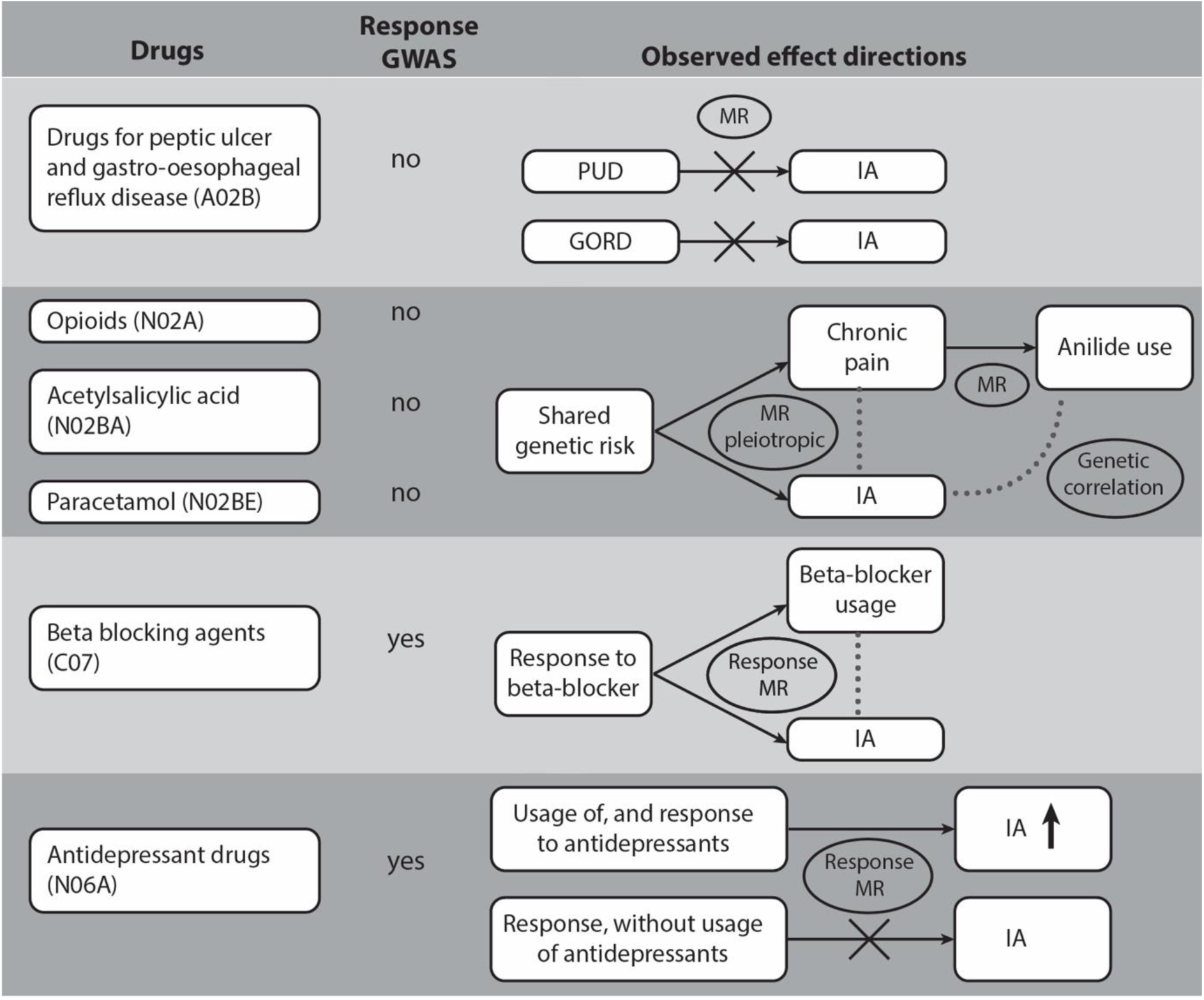
The relationships between intracranial aneurysms (IA) and the drug usage traits correlated with IA. Usage of drugs from six classes (column 1) was correlated with IA. The second column shows the existence (+) or absence (0) of a genetic correlation of the drug class with intracranial aneurysms (IA), ischemic stroke (IS) and intracerebral hemorrhage (ICH). Column 3 indicates whether a drug response GWAS was available for a drug or drugs within the class. Column 4 shows the relationships between genetically predicted drug class usage, drug indications, drug response, and IA.

## Discussion

We identified genetic correlations between IA usage of drug classes antidepressant drugs, paracetamol, acetylsalicylic acid, opioid drugs, beta-blockers and drugs for peptic ulcer and gastro-oesophageal reflux disease. For the anti-depressant drugs, we found evidence that its usage influences the risk of IA, although it involves a risk-increasing effect rather than a risk-reducing one. The other observed correlations can be explained by pleiotropic relationships between IA, the drug indication and drug usage (e.g., between IA, chronic pain, and pain medication usage). However, based on our results we cannot provide sufficient evidence that these studied drug classes directly influence the risk of IA and the exact reason for the observed pleiotropic relationships remains to be investigated.

We found a potential link between antidepressant drug usage and liability to IA and ASAH. Our findings may imply an adverse response to antidepressant drugs. Depression has been linked to stroke liability in women in epidemiological studies. ^28^ Since depression is more prevalent in women it may be a factor explaining why the incidence of ASAH is higher in women. The sex-specificity of depression as a risk factor for IA has not yet been studied. Inferring a causal adverse response should be done with caution for several reasons. First, we were unable to identify the drug(s) within the antidepressant drug class that explain the observed increase in IA and ASAH liability, which is an important follow-up step. Second, in theory, responsiveness to antidepressant drugs may instead be explained by a (hidden or unknown) type or cause of depression which in turn may affect IA and ASAH risk. No evidence for such mechanisms currently exists. A prospective cohort study of the effect of depression and antidepressant drug use on IA or ASAH could provide further insight in our findings.

We found a novel pleiotropic link between genetically predicted paracetamol usage and liability to IA. Based on our genetic correlation analysis, liability to the usage of paracetamol is associated with an increased risk of IA. Paracetamol has been proposed to lower body temperature and thereby protect from severe outcome of other stroke types being acute ischemic and hemorrhagic stroke, but this is not yet supported by evidence from a large clinical trial, and no evidence for a role in ASAH has been shown. ^29^ Further studies to identify the causal genes underlying the observed pleiotropy between paracetamol and IA may help understand the underlying pathogenic processes.

A previous study investigated the causality of the same 23 drug usage traits on various stroke types, including a smaller GWAS of IA with participants from Finland. ^30^ The authors found a similar seemingly risk-increasing effect of liability for the usage of cardiovascular system drugs (diuretics, beta-blockers, and renin-angiotensin system drugs) on ASAH. Given that these findings imply that drugs used to treat cardiovascular disease would increase cardiovascular disease risk these observations are likely driven by pleiotropy and should be interpreted with great caution. In our study which included the necessary downstream and sensitivity analyses we found no evidence for a direct effect or side-effect of cardiovascular system (anatomical therapeutic chemical code C) drug class usage on IA.

Strengths of this study include the use of large GWAS summary statistics for IA and drug usage, the comprehensiveness of our approach, and the use of multiple MR methods to validate our findings. We performed thorough attempts to deal with pleiotropy and potential reverse causation. The CAUSE MR method was previously shown to be most robust to confounding by estimating confounders among several MR methods. ^17^ However, in our follow-up analyses were showed that BP was in fact the factor driving the genetic overlap between usage of beta-blockers and liability to IA. In the Supplementary Data we provide a detailed discussion of limitations: limits of extrapolating the results, incomplete correction for BP, unbalanced representation of drugs within a class, and correlation between drug classes.

In conclusion, we found evidence for a risk-increasing effect of drugs within the anti-depressant drug class on IA, which effect should be further explored in prospective cohort studies. Lastly, for paracetamol, acetylsalicylic acid, opioid drugs, and drugs for peptic ulcer disease and gastro-oesophageal reflux disease, and beta-blockers, we found shared genetic risk underlying IA. Future studies aiming to untangle these shared mechanisms may improve our understanding of the pathogenesis of IA and ASAH, which may in turn identify processes that can be perturbed to affect the liability to IA and ASAH.

Supplementary data references: ^31^ ^32^

## Data Availability

Only publicly available aggregated data was used with informed consent in place in the original studies. Data generated in this study is available in the Supplement.

## Acknowledgements

We thank the international stroke genetic consortium IA working group for providing GWAS summary statistics for IA.

This work was undertaken under UK Biobank project number 2532.

## Non-standard Abbreviations and Acronyms

ASAH: aneurysmal subarachnoid hemorrhage
BP: blood pressure
CAUSE: Causal Analysis Using Summary Effect Estimates
CI: confidence interval
CMP: Chronic multisite pain
*CNNM2*: Cyclin and CBS domain divalent metal cation transport mediator 2
GSMR: generalized summary statistics-based Mendelian randomization
GWAS: genome-wide association study
IA: intracranial aneurysm
MR: Mendelian randomization
OR: odds ratio
PGS: polygenetic score
ρ_g_: genetic correlation
SSRI: selective serotonin reuptake inhibitor

## Supplemental Material

Detailed Methods

Supplementary Figures 1-7

Supplementary Tables 1-9

References 31-32

STROBE-MR checklist

